# Nutritional risk assessment using Malnutrition Universal Screening Tool (MUST) in COVID-19 patients: An observational study in a tertiary care hospital in Eastern India

**DOI:** 10.1101/2021.07.21.21260904

**Authors:** Sudeshna Maitra Nag, Subhrojyoti Bhowmick, Sayantani Bhowmick, Uttiya Deb, Debarati Kundu, Krishnangshu Ray, Sujit Kar Purkayastha

## Abstract

**Aims:** To diagnose malnutrition, the nutritional status of each infected patient should be evaluated before starting general treatment. The role of Malnutrition Universal Screening Tool (MUST) in evaluating nutritional status of COVID-19 patients is still unknown. The aim of this study was to evaluate the use of MUST in assessment of nutritional status of COVID-19 patients.

**Methods:** We retrospectively analyzed the data of hospitalized COVID-19 patients above 18 years of age from July 25^th^ to September 25^th^, 2020. All COVID-19 patients with a length of hospital stay greater than 24 hours underwent malnutrition screening and nutritional assessment based upon MUST. Demographic data, laboratory parameters and MUST score were retrieved from case files.

**Results:** Out of 106 COVID-19 patients included in the study, 68 (64%) were male and 38 (36%) were female. Number of deaths due to COVID-19 was 17 (16.03%). A total of 22 (20.75%) patients had MUST score of 2 and above. Analysis between MUST score and age group showed statistically significant result (p=0.012). MUST score according to clinical outcome at the end of hospitalization was also statistically significant (p<0.001).

**Conclusion:** Our results highlight a possible role of MUST as screening tool for malnutrition in COVID-19 patients.

## Introduction

COVID-19 is a newly diagnosed disease which was first identified in late December 2019 in Wuhan, China.^[1]^ It is a highly contagious disease that transmits via respiratory droplets, aerosol and mucosal membrane contact with fomites.^[2,3]^ World Health Organization (WHO) declared the COVID-19 outbreak a pandemic on 11th March 2020.^[4]^

Malnutrition is one of the key reasons due to which human immune system is severely affected.^[5]^ Reduced food intake, which increases malnutrition risk in COVID-19 patients, is caused by nausea, diarrhea and loss of appetite.^[6]^ It has been observed that malnutrition prolongs hospitalization periods.^[7]^ Diagnosis and management of malnutrition, therefore, must be included in management of hospitalized COVID-19 patients.^[8]^ To diagnose malnutrition, the nutritional status of each infected patient should be evaluated before starting general treatment.^[9]^ Some of the pre-validated recommended tools for screening nutritional risk are Nutritional Risk Screening 2002 (NRS 2002) score,^[10]^ Nutrition Risk in the Critically ill (NUTRIC) score ^[11,12]^ and Malnutrition Universal Screening Tool (MUST).^[13]^

Due to the emergence of novel coronavirus, there is a paucity of data on nutritional status of COVID-19 patients. It is also unknown whether MUST scoring is applicable in this scenario. Therefore, this retrospective observational study was conducted to evaluate the use of MUST in assessment of nutritional status of COVID-19 patients.

## Materials and methods

This retrospective observational study was conducted in a tertiary care hospital in Eastern India from July 25 to September 25, 2020. The institutional ethics committee approved the proposal of this study. The diagnosis of COVID-19 positive patients was done based on the following:

i. Nasopharyngeal and oropharyngeal swab test
ii. History of epidemiological exposure
iii. Clinical symptoms such as fever, cough, respiratory distress and gastroenterological symptoms, and
iv. Pulmonary imaging

### Study population

As our hospital was previously a non-COVID hospital, the hospitalized patients, the staffs including physicians, nurses and other operating personnel with symptoms or with history of contact with COVID-19 positive patients were tested for COVID-19 and thereafter admitted if found to be COVID-19 positive. Therefore, the study population was both critically-ill and non-critical patients. Patients were excluded from the study if they were: 1) <18 years; 2) pregnant; 3) had a length of hospital stay <24 hours.

### Data collection

All COVID-19 positive patients with a length of hospital stay of more than 24 hours underwent malnutrition screening and nutritional assessment based upon the Malnutrition Universal Screen Tool (MUST) by a team of assigned dietitians. The MUST score is a prevalidated tool developed by the Malnutrition Advisory Group of the British Association of Parenteral and Enteral Nutrition (BAPEN) for use in all health care settings.^[13]^ MUST is designed to identify need for nutritional treatment as well as establishing nutritional risk based on the association between impaired nutritional status and function. It includes body mass index (BMI) score, a weight loss score and an acute disease score.^[14]^ Clinical findings and other relevant investigational data were retrieved from the case files of the COVID-19 patients for data analysis.

### Statistical Analysis

Descriptive statistics was used to analyze the data. Results were expressed as number of patients, percentage(%), mean and standard deviation(SD). Categorical variables were compared using Pearson’s chi-squared test. P<0.05 was considered as statistically significant. Statistical analysis was done using IBM SPSS 21.0 software and Microsoft Office Excel 2007.

## Results

A total of 106 COVID-19 positive patients were included in the study. A summary of demographic characteristics of our study participants is reported in Table 1. Among them 68 (64%) were male and 38 (36%) were female. A total of 22 patients (20.7%) belonged to 18-30 years age group, 31 patients belonged to 30-50 years age group (29.2%), and the remaining above 50 years of age (Table 1). The mean age of the patients was 47.3 years. The patients had mean (± SD) duration of hospital stay of 14.9 ± 8.7 days (Table 1). The initial symptoms reported were fever (46.2%), dyspnea (35.8%) and cough (34.9%). Our study population also consisted of asymptomatic COVID-19 patients (12.3%). The most associated co-morbidity in our study participants was diabetes (24.5%) followed by chronic obstructive pulmonary disease (17.9%) and chronic kidney disease (9.4%) as shown in Table 1. The mean (± SD) hemoglobin level was 12.1 ± 2.28 g/dl (Table 2). The mean (± SD) serum albumin level was 3.30 ± 0.88 g/dl (Table 2).

**Table 1.**
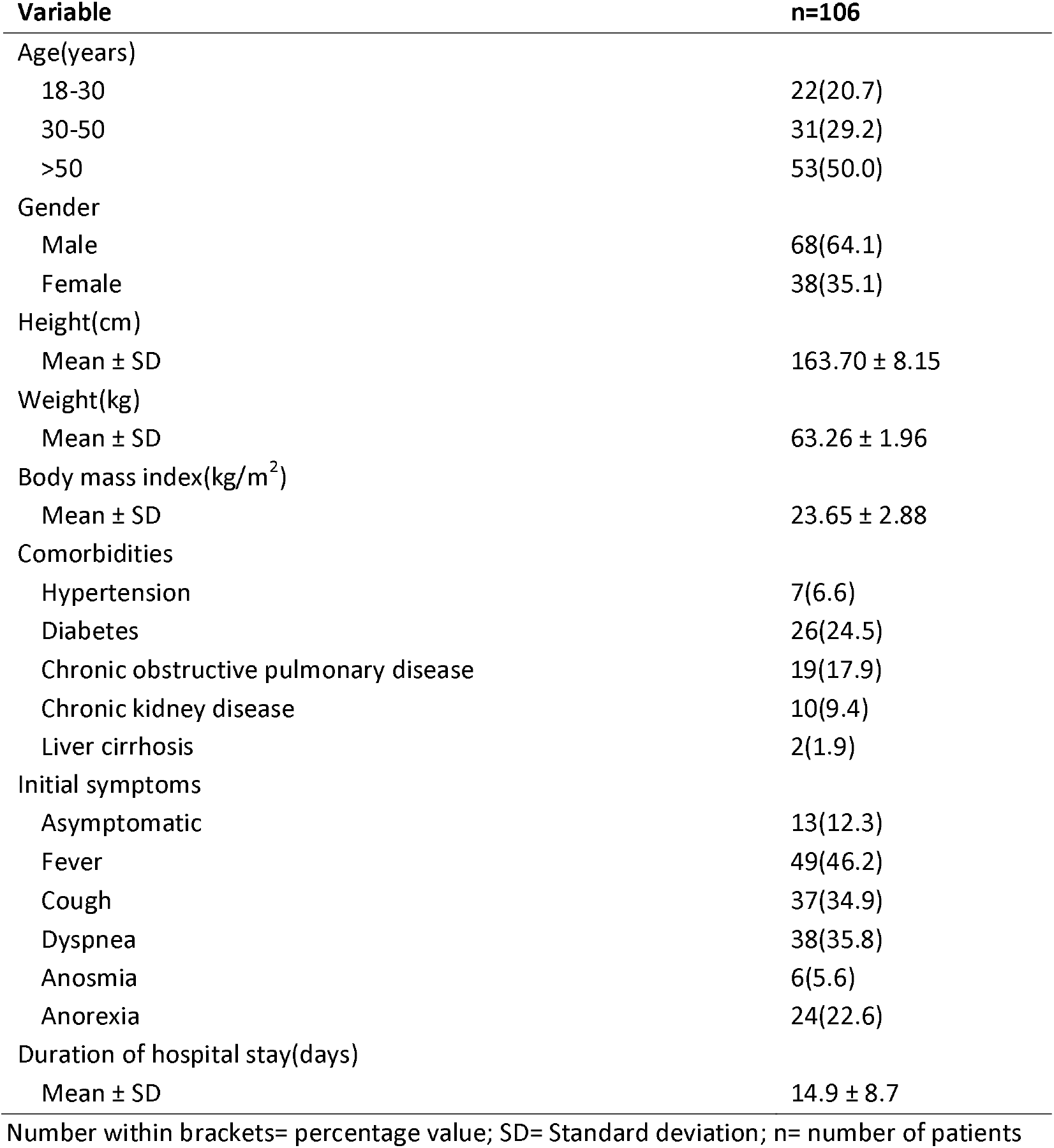
Demographic characteristics of study participants.

**Table 2.** Laboratory parameters of study participants.

| **Variable** | **Mean ± SD** |
| --- | --- |
| Haemoglobin(g/dl) | 12.1 ± 2.28 |
| Total leukocyte count(/cu.mm) | 8208.1 ± 5745.68 |
| Total protein(g/dl) | 6.96 ± 1.02 |
| Serum albumin(g/dl) | 3.30 ± 0.88 |
| AST(IU/l) | 52.9 ± 30.8 |
| ALT(IU/l) | 54.8 ± 43.7 |
| Serum sodium(mEq/l) | 137 ± 4.91 |
| Serum potassium(mEq/l) | 4.1 ± 0.67 |
| Serum creatinine(mEq/l) | 1.51 ± 2.65 |

AST= Aspartate aminotransferase; ALT = Alanine aminotransferase

Among 106 patients, 89(83.97%) patients were discharged from the COVID-19 ward. A total of 17 deaths (16.03%) were reported during our study period (Table 3). No deaths occurred in 18-30 years age group. The maximum number of deaths (14) was observed in above 50 years of age group followed by 3 deaths in 30-50 years age group. A summary of clinical outcome (death or discharge) according to age group is reported in Table 3.

**Table 3.** Clinical outcome of the study participants according to age group.

| **Clinical outcome** | **n(%)** |
| --- | --- |
| Discharged |  |
| 18-30 | 21(19.8) |
| 30-50 | 34(32.0) |
| >50 | 34(32.0) |
| Deaths |  |
| 18-30 | 0(0.0) |
| 30-50 | 3(0.03) |
| >50 | 14(0.21) |

n=number of patients

MUST scoring was done for all patients during nutritional assessment. A total of 22 (20.75%) patients were found to have MUST score of 2 and above indicating a high risk of malnutrition followed by 13(12.26%) patients having MUST score 1 and remaining having MUST score 0. Out of 17 deaths due to COVID-19, 11(64.7%) patients had MUST score of 2 and above (Figure 3). When we did analysis between MUST score and age group of the patients, the results were statistically significant (p=0.012) (Figure 2). Analysis of MUST score according to clinical outcome (death or discharge) at the end of hospitalization also showed statistically significant result (p<0.001) (Figure 3).

**Figure 1:**
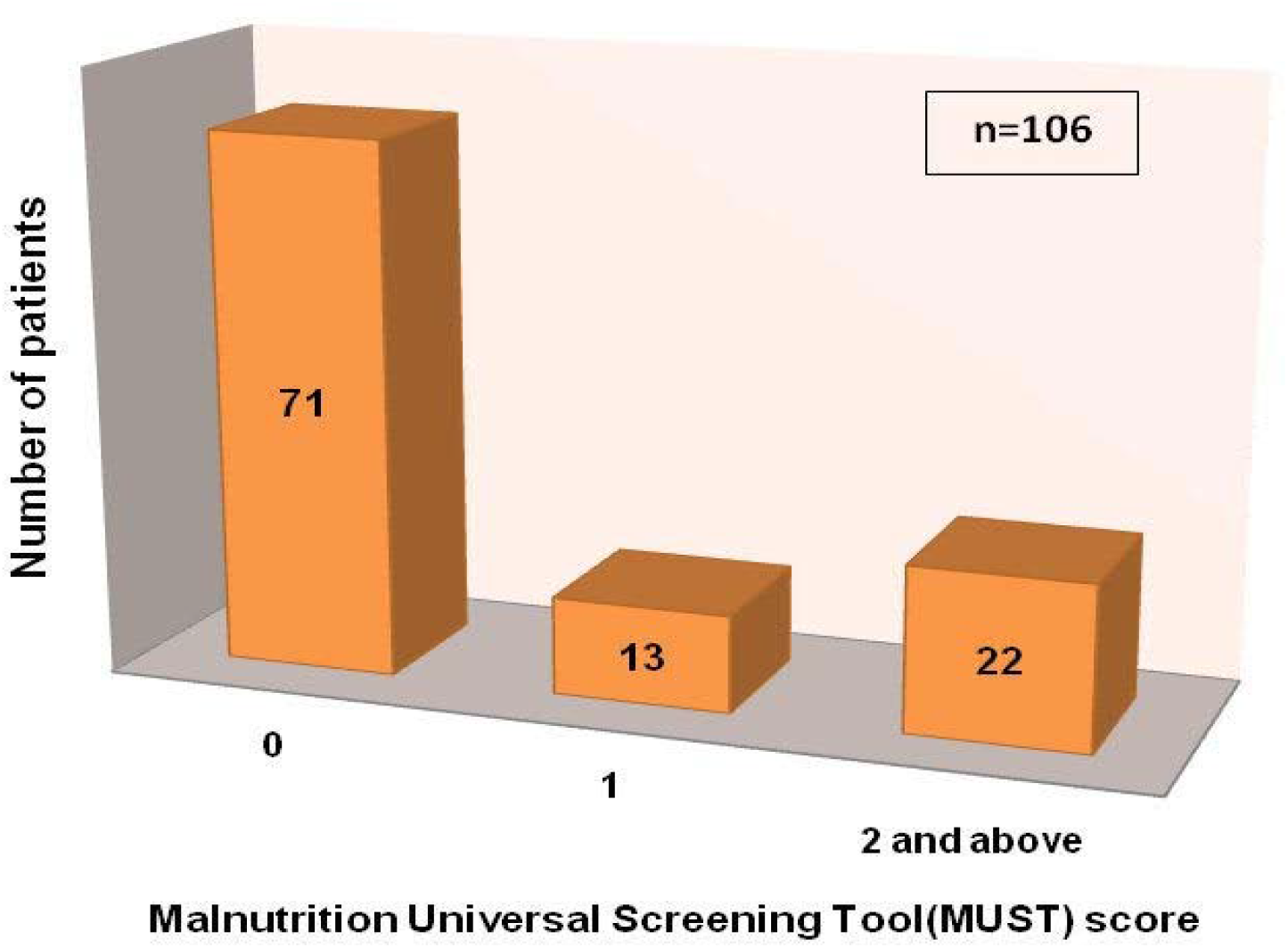
Malnutrition Universal Screening Tool (MUST) score

**Figure 2:**
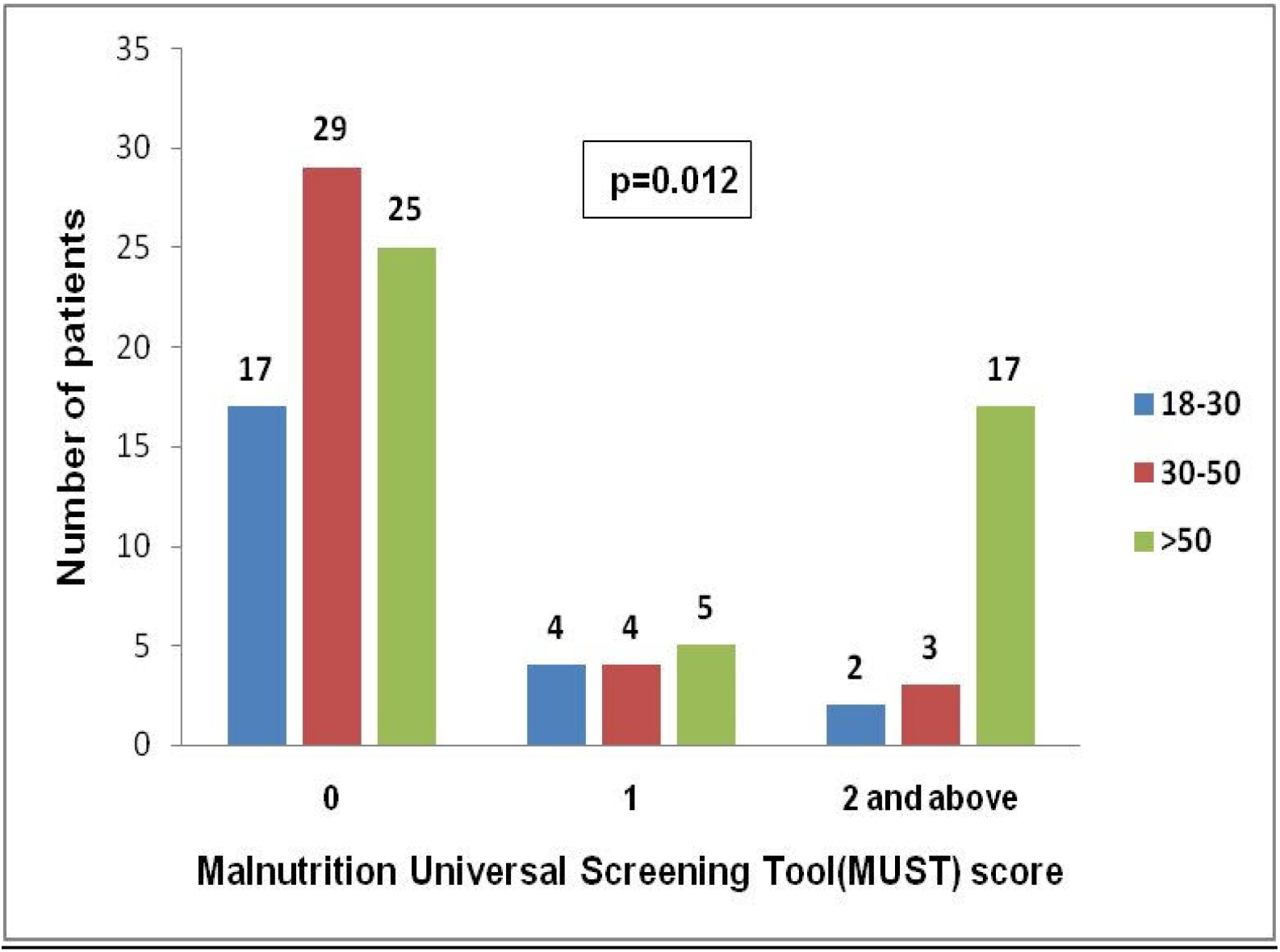
Malnutrition Universal Screening Tool (MUST) score according to age group

**Figure 3:**
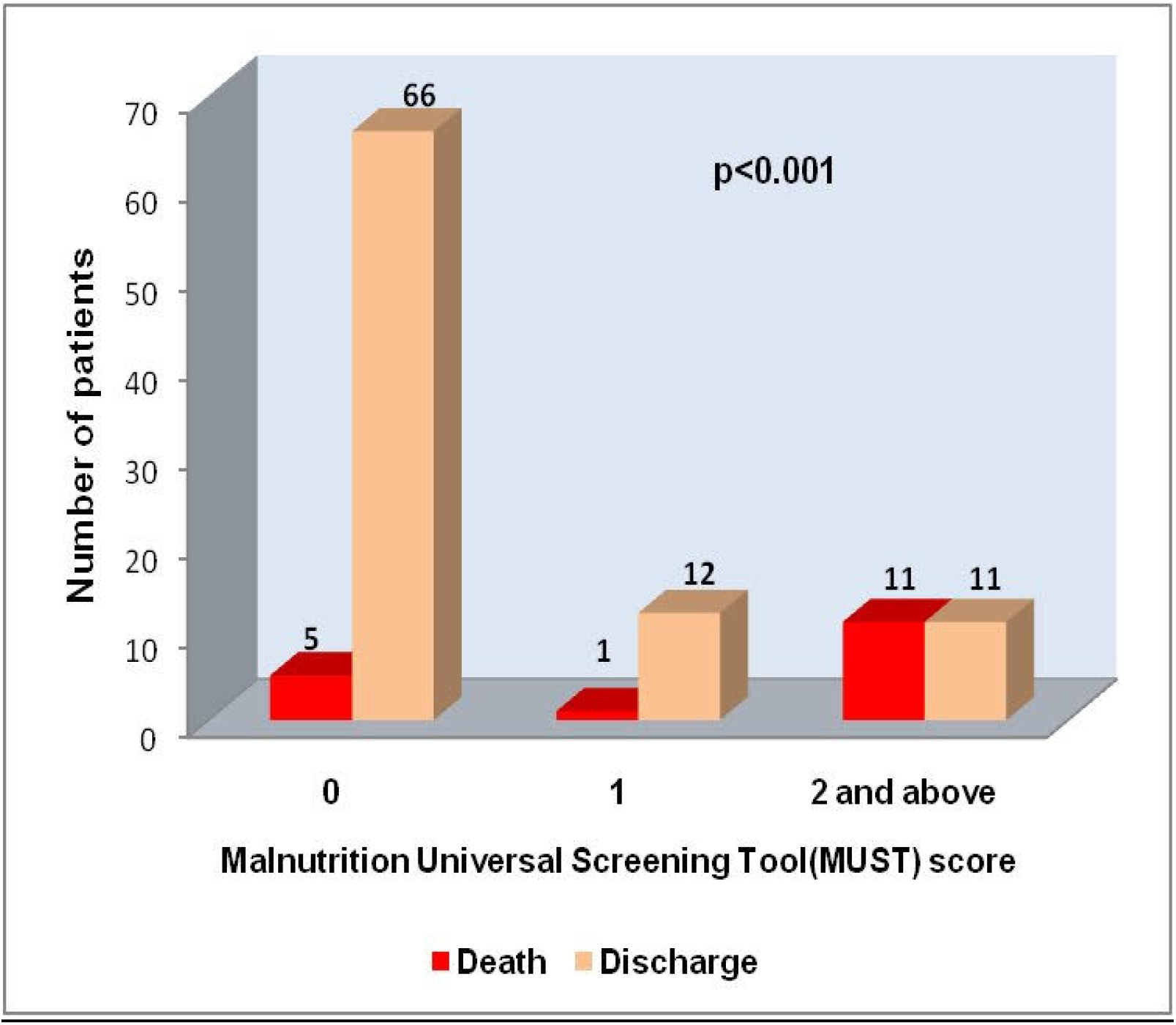
Malnutrition Universal Screening Tool (MUST) score according to clinical outcome

## Discussion

Malnutrition is known to be prevalent in hospitalized patients.^[15]^ It causes adverse effects on physical and psychological function.^[16]^ Therefore, routine screening of nutritional status is recommended in all patients admitted to hospital to allow early intervention.^[17]^ MUST tool can easily identify risk of malnutrition in all hospital settings.^[13]^ Correct identification of these patients, who are at greater need of nutritional intervention, allows appropriate targeting of otherwise scarce dietetic expertise and resources in a developing country like India. The applicability of MUST for COVID-19 patients is largely unknown. In this study, we for the first time report the use of MUST for assessment of nutritional status in COVID-19 patients.

Till date the accuracy of MUST to identify critical patients under nutrition risk is not well-established.^[13]^ In this study, we could identify the patients with high nutritional risk. Most of the patients with high nutritional risk were above 50 years of age. The mortality was also high among these patients. Russel et al. reported that older hospitalized patients were more likely to be malnourished than younger people based on MUST scores.^[18]^ One single centre study examined the ability of MUST to predict mortality and length of stay in older people.^[19]^ But it is not clear whether a high nutrition risk score was an independent predictor of mortality or not. According to a study done on critically ill COVID-19 patients in a hospital in Wuhan, China where NUTRIC score was used to assess the nutritional status of the patients, the high nutritional risk group demonstrated significantly higher mortality of ICU 28-day as well as twice the probability of death at ICU 28-day than the low nutritional risk group.^[20]^ But till date there is limited evidence of the effectiveness of MUST in COVID-19 patients.

The advantage of our study is that our hospital was not a designated COVID hospital, so there was no bias in patient selection. Each and every patient got admitted as soon as they tested positive for COVID-19. Our patients ranged from being asymptomatic carriers to being critically-ill. Also, we used MUST for both critical and non-critical COVID-19 patients. This gives novelty to our work.

The limitations of this study are small sample size and retrospective observational study design. Randomized controlled studies with larger sample sizes are required to determine whether MUST can effectively assess nutritional risk in COVID-19 patients.

## Conclusion

In summary, our study is the first to investigate the use of MUST score for assessing nutritional risk in COVID-19 patients. Based on MUST score at admission, a high nutritional risk was observed in elder patients. Higher MUST score was also associated with higher mortality. Therefore, the MUST score may be an appropriate tool for nutritional risk assessment in COVID-19 patients; however, studies with larger sample sizes are needed before generalization of the study findings in COVID-19 patient care.

## Data Availability

All data archived

